# Home energy efficiency, overcrowding and lower respiratory tract infection admissions in infants: national birth cohort study in Scotland

**DOI:** 10.64898/2026.08.19.26360387

**Authors:** Caroline Hart, Amal Rammah, Marianna Riccio, Bianca De Stavola, Jonathon Taylor, Phil Symonds, Steve Cunningham, Chris Dibben, Olivia Swann, Samantha Hajna, Pia Hardelid

**Affiliations:** Environmental Child Health Research Group, UCL Great Ormond Street Institute of Child Health, London, United Kingdom; Department of Infectious Diseases and Public Health, University of Rome La Sapienza, Rome, Italy; Department of Civil Engineering, Tampere University, Tampere, Finland; Institute of Environmental Design and Engineering, University College London, London, UK; Department of Child Life and Health, Institute of Regeneration and Repair, University of Edinburgh, Edinburgh, UK; School of Geosciences, University of Edinburgh, Edinburgh, UK; Usher Institute, University of Edinburgh, Edinburgh, UK; Department of Health Sciences, Brock University, St Catharines, Ontario, Canada

## Abstract

**Background:** We examined whether two key housing quality indicators, energy efficiency and household overcrowding, were associated with lower respiratory tract infection (LRTI) hospital admissions in infants.

**Methods:** We used a cohort of all singleton births in Scotland 2010-2012, created through linked vital statistics and health data. LRTI admissions were characterised in hospital records. Overcrowding (defined using the national room standard) and median postcode-level energy efficiency were defined using maternal Census and postcode-level Energy Performance Certificate data linked to the cohort, respectively. We used logistic regression to model the odds of at least one infant LRTI admission.

**Results:** The cohort included 136,123 infants of whom 4.0% had at least one LRTI admission. Overcrowding was more common among infants of younger mothers and those in rented housing. Energy efficiency was lower among infants of older mothers, living in owner occupied homes, in less deprived areas. Compared with infants living in homes with excess rooms (under-occupied housing), those whose homes were below, or met, the minimum room standard had higher odds of LRTI admission (adjusted odds ratio 1.07, 95% CI 0.98–1.17; 1.10, 95% CI 1.03–1.17, respectively). Postcode-level energy efficiency was not associated with LRTI admission odds.

**Conclusion:** Overcrowding was more common in socioeconomically disadvantaged households and associated with increased risk of LRTI admission in infancy. Lower energy efficiency was associated with factors commonly linked to socioeconomic advantage and was not associated with LRTI admissions. Improving access to housing with adequate living space may reduce the burden of LRTIs in early life.

**Key messages:** xxxxx

## Background

Lower respiratory tract infections (LRTIs), including bronchiolitis, bronchitis, and influenza are a common reason for hospital admissions among infants in the UK and globally[1]. Although viral LRTIs are frequently self-limiting, they may also cause severe respiratory symptoms, particularly among infants, and children born prematurely or with underlying chronic conditions[2]. Early-life LRTIs have been associated with recurrent wheeze, asthma and respiratory mortality[3, 4].

- What is already known on this topic: Housing conditions, including crowding and property energy efficiency, have been associated with childhood respiratory tract infection admission risk.
- What this study adds: Using a whole-nation birth cohort in Scotland, we found that residential crowding and high energy efficiency was more common among infants whose mothers were younger, and living in poorer areas. Infants whose homes were below, or met, the minimum room standard (measure of crowding) had a higher risk of respiratory tract infection admission; energy efficiency was not associated with the risk of admission.
- How this study might affect research, practice or policy: Ensuring that families with young children have access to housing with living space proportionate to family size may help reduce the burden of LRTIs in infancy.

Studies in multiple countries have found that LRTI hospital admission rates are highest in socio-economically deprived areas, and in families experiencing poverty. Several socio-demographic and environmental factors, including environmental tobacco smoke and ambient air pollution exposure, number of older siblings and daycare attendance may explain some of these inequalities[5–7].

Poor housing conditions may be another pathway linking socioeconomic deprivation to LRTIs. Young children can spend up to 80% of their time in the home[8], and are more vulnerable to indoor environmental exposures, as their lungs and immune systems are still developing[9]. Infants and preschool children are therefore more susceptible to the adverse effects of poor-quality housing including overcrowding, poor indoor air quality, damp or mould. Previous studies suggest children residing in crowded[10] or damp/mouldy housing[11] are at a higher risk of LRTIs and associated hospital admissions. Energy efficiency of the home is a key determinant of indoor air quality[12] and is strongly associated with the risk of dampness and mould growth[13], due to the higher expense required to heat energy inefficient homes, increasing the risk of damp and condensation. Energy efficient, well insulated homes can be more airtight, which without adequate supplementary ventilation enhances the spread of respiratory viruses[14] and may increase levels of indoor-generated air pollutants. Airtightness can also limit the infiltration of outdoor air pollutants[14]. An ecological study in England, found that areas with higher rates home energy efficiency improvements had higher rates of adult respiratory and cardiovascular hospital admission[15]. They hypothesised homes with energy efficiency interventions without adequate ventilation may have worse air quality.

Compared to other European countries, the UK housing stock is generally older (one in five UK properties are over 100 years old) and poorly insulated with lower energy efficiency[16]. Further, 12.7% of families with children in the UK live in overcrowded housing[17]. The UK government has recently introduced several new laws and initiatives to improve housing[18] [19, 20] [21]. Therefore, our aim was to study the associations of overcrowding and energy efficiency, known to be socially patterned, with the risk of having one or more LRTI admissions during infancy.

## Methods

### Data sources

We used a linked national birth cohort of all children born in Scotland between 2010 and 2012, including vital statistics, health, Census and housing data. The cohort was derived for an analysis previously published and has been described in detail elsewhere[22]. Briefly, the spine of the birth cohort included birth records for all liveborn children in Scotland between 2010 and 2012, linked to mortality records, maternal 2011 Census records, maternity records (Scottish Morbidity Records [SMR]-02) and hospital admission records (SMR-01). Data were linked using the Community Health Index (CHI) number, a unique personal identifier used in the Scottish National Health Service (NHS), by Public Health Scotland. The CHI Register (the Scottish NHS national address register) was used to link to postcode-level data on home energy efficiency ratings from Energy Performance Certificates (EPCs). Appendix Table A1 summarises all datasets and the variables extracted from each for this study.

The EPCs constitute a dataset holding information on the energy efficiency of all homes sold or rented to new tenants since 2008, and any new buildings constructed since 2013.[23]. EPC records provide information directly observed by property surveyors about building characteristics, including insulation, heating fuel type and floor space, and an estimate of building energy efficiency. The certificates are valid for up to 10 years; however, renewing them is non-mandatory, unless there is a change of tenancy/ownership. The EPC data used in this study were extracted in 2021. As multiple certificates can exist for one dwelling, only the most recent certificate record for each dwelling was extracted. To ensure as complete data for as many postcodes as possible, we included EPCs from any time until 2021. We derived the median energy efficiency rating at each postcode. We selected a child’s first recorded postcode on the CHI register, assumed to be the residential postcode at birth, and used this to link to the EPC register. The small number of children living in postcodes at birth without at least one EPC were excluded.

### Study population and period

Our study population consisted of singleton children born to Scottish resident mothers between 1^st^ January 2010 and 31^st^ December 2012. Births recorded as <24 weeks’ gestation or with a birth weight <500g were excluded (indicative of stillbirths), as were children who died or emigrated from Scotland before their second birthday. All children without a linked maternal census record were excluded, and children who did not live in private households (eg children living in communal housing such as hostels according to Census data) or were living in rent-free housing were also excluded as we would have been unable to assign exposures to these children. When linking children’s health data to EPC records, we excluded infants missing a postcode record in the CHI register, those with no EPC records in their postcode, and those for whom the first postcode timestamp occurred after their first birthday.

### Outcome

Our outcome was characterised as one or more LRTI hospital admissions in the first year of life (yes/no) using linked hospital admission (SMR-01) data. LRTI admissions were defined as any hospital admissions where an acute LRTI was recorded as the primary diagnoses using International Classification of Diseases, 10th edition (ICD- 10), diagnostic codes J10- J22[22].

### Exposures

#### Energy efficiency

Energy efficiency ratings were obtained from EPC records. The ratings are adjusted for floor size and expressed on a scale of 1-100, where a rating of 1 is indicative of very poor energy efficiency and expensive heating costs and 100 is indicative of very high energy efficiency and low heating costs. In rare cases, the rating may exceed 100, in which case the dwelling is generating more energy than it is using. Ratings are categorized into bands A-G (Appendix Table A2). We dichotomised the median postcode-level EPC values as high efficiency (≥69; 69 is the median EPC rating in Great Britain[24, 25]), which equates to a high EPC rating (A-C)) or low efficiency (<69, which equated to a poor energy efficiency rating (D-G). As not all properties in a postcode had an EPC, we also calculated the ratio of EPCs per number of properties by postcode in 2011.

#### Residential overcrowding

The National Records for Scotland derived an ‘occupancy rating’ variable from data collected in the 2011 maternal Census by subtracting the notional number of rooms required (determined based on the number of available rooms in relation to number of adults and the number, ages and sex of children[26]). We classified crowding using this variable as follows: >minimum room standard (at least 1 room more than the minimum (basic) room standard); =minimum room standard (meets the minimum standard); and <minimum room standard (at least 1 room less than the minimum standard). We refer to the category <minimum room standard as overcrowded and > minimum room standard as under-occupied. Note that the number of rooms, not the number of bedrooms, was used in the 2011 Census to measure overcrowding (see discussion).

#### Composite adverse housing measure

We further derived a composite measure for adverse housing conditions by combining the overcrowding and energy efficiency variables as follows: 1) no adverse housing conditions (energy efficiency rating ≥69 and having at least the basic room standard); 2) one adverse housing condition only (either energy efficiency rating <69 or <basic room standard); and 3) two adverse housing conditions (both energy efficiency rating <69 and less than the basic room standard).

#### Covariates

The study population was described according to the following child-level characteristics: biological sex (recorded at birth), dichotomised season of birth (spring/summer vs. autumn/winter), gestational age (preterm, <37 weeks; term, 37- 41 weeks; late term, 41+ weeks), and birthweight (<2,500/2,500-3,500/>3,500 grammes). Family socioeconomic position was characterised by maternal highest qualification level (<Level 2 versus ≥Level 2, where Level 2 is Scottish Certificate of Education–advanced or equivalent, for which national exams taken at age 18 years),[27] maternal age (<20/20-29/30-39/<u>≥</u>40 years), maternal country of birth (UK/ outside UK) and housing tenure (owner occupied/social rented/private rented).

We also considered residential and neighbourhood characteristics. Urban/rural status of children’s residential addresses from birth records was assigned using the 8-fold Scottish Government urban/rural classification[28], where towns/cities with a population≥3000 were considered urban. Accommodation type was derived from the Census and categorised as detached, semi-detached, terraced, or flat/maisonette. A small number of children who lived in mobile homes or temporary accommodation were assigned to the flat/maisonette group to avoid individual disclosure. Heating type was also derived from the 2011 Census and grouped as gas central heating, solid fuel heating, no central heating, or other heating type (electric, storage heaters, oil). Quintiles of the Scottish Index of Multiple Deprivation (SIMD), a small-area-level indicator of deprivation for Data Zone areas (average population of 500-1,000 people), were derived from the address on the birth record.

#### Statistical methods

For each variable of interest, we described its distribution among children in the cohort. We examined how postcode-level energy efficiency and residential overcrowding varied among children according to child, family and residential/neighbourhood characteristics.

We fitted logistic regression models to separately examine the associations between postcode-level energy efficiency categories (A-C versus D-G) and residential overcrowding and one or more infant LRTI admissions. In energy efficiency models, a high postcode-level energy efficiency rating (A-C) was taken as the reference group. Having more than the minimum required rooms was the reference group in the residential crowding models. The adjustment variables included in the multivariable models were selected based on a directed acyclic graph (DAG) developed using DAGitty[29] (Appendix figures A1 and A2) for the presumed relationships among the variables of interest. The energy efficiency model was adjusted for area of residence (urban/rural), maternal age, maternal qualifications, housing tenure, maternal country of origin, accommodation type and heating type. The residential overcrowding model was adjusted for area of residence, maternal age, maternal highest qualification, housing tenure, maternal country of origin and accommodation type. Results were expressed as crude and adjusted odds ratios (OR) with 95% confidence intervals. Only children with complete data for all the adjustment variables were included in our analyses.

## Results

Our cohort included 136,123 children (Figure 1). The proportion of children missing either a postcode reference record, EPC record, or a first postcode record <12 months of age was low (0.01 %, 0.5% and 0.8%, respectively). The mean ratio of number of EPC records to properties by postcode in 2011 was 0.55. 5,491 cohort children (4.0%) had at least one LRTI admission during infancy. A total of 74,627 children (54.8%) resided in a postcode at birth with an energy efficiency at or above the median value (68.0), and 18,661 children (13.7%) lived in overcrowded households. Less than 5% of children had missing data on any single variable (Appendix Table A3). The distributions of the variables of interest were similar in the in the analytical cohort and in the full cohort (Appendix Table A3; Table 1).

**Figure 1.**
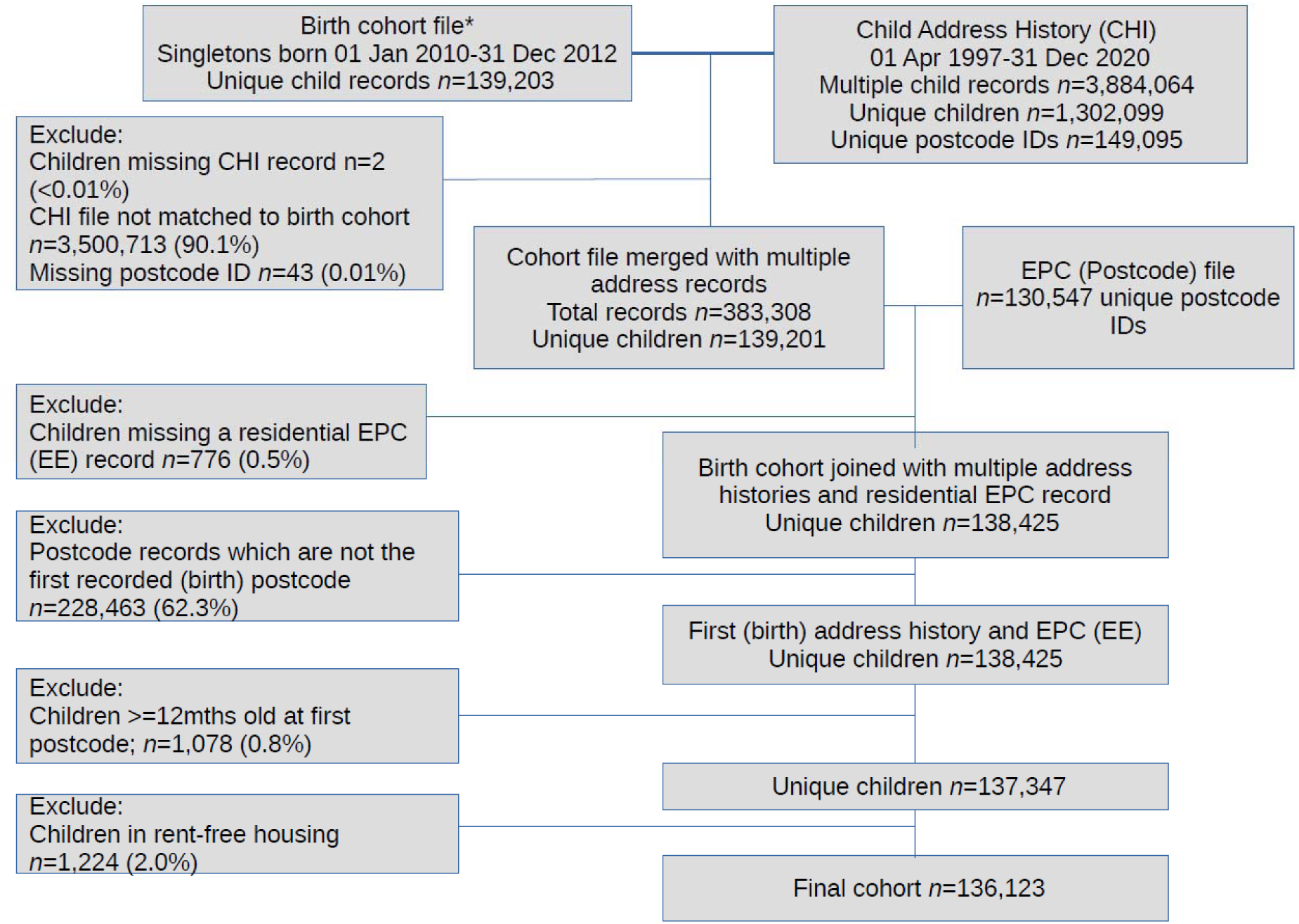
Flow diagram of linkages and exclusions to develop the birth cohort, Scotland, 2010-2012. *Total singleton births in Scotland to Scottish resident mothers 2010-2012: 169,706. The birth cohort file referred to here also excluded children who died or migrated out of Scotland before their 2 birthday, living in communal dwellings, or who were born at <24 weeks’ gestation or with a birthweight of <500g (n=2,905) [22].

**Table 1.** Number and proportion of infants by postcode-level energy efficiency according to child, household and neighbourhood characteristics, Scotland, 2010- 2012. N=132,665.

|  | Overall | Postcode-level energy efficiency |  |
| --- | --- | --- | --- |
| Characteristic |  | High | Low |
| **Sex** |  |  |  |
| Male | 67946 (51.2) | 30706 (45.2) | 37240 (54.8) |
| Female | 64719 (48.8) | 29398 (45.4) | 35321 (54.6) |
| **Season of birth** |  |  |  |
| Spring/summer | 66657 (50.2) | 30184 (45.3) | 36473 (54.7) |
| Autumn/winter | 66008 (49.8) | 29920 (45.3) | 36088 (54.7) |
| **Gestational age** |  |  |  |
| Preterm | 7044 (5.3) | 3278 (46.5) | 3766 (53.5) |
| Term | 91676 (69.1) | 41740 (45.5) | 49936 (54.5) |
| Late term or missing<sup>a</sup> | 33945 (25.6) | 15086 (44.4) | 18859 (55.6) |
| **Birth weight** |  |  |  |
| <2,500g | 6116 (4.6) | 2932 (47.9) | 3184 (52.1) |
| 2,500-3500g | 64145 (48.4) | 29293 (45.7) | 34852 (54.3) |
| >3,500g or missing<sup>a</sup> | 62404 (47) | 27879 (44.7) | 34525 (55.3) |
| **Maternal level of education** |  |  |  |
| Less than level 2/not applicable* | 47066 (35.5) | 22856 (48.6) | 24210 (51.4) |
| Level 2 or higher | 85599 (64.5) | 37248 (43.5) | 48351 (56.5) |
| **Maternal age group** |  |  |  |
| <20 years | 7471 (5.6) | 3549 (47.5) | 3922 (52.5) |
| 20-29 years | 58623 (44.2) | 28063 (47.9) | 30560 (52.1) |
| 30-39 years | 61531 (46.4) | 26668 (43.3) | 34863 (56.7) |
| ≥40 years | 5040 (3.8) | 1824 (36.2) | 3216 (63.8) |
| **Marital status** |  |  |  |
| Married/cohabitating | 111542 (84.1) | 49619 (44.5) | 61923 (55.5) |
| Separated/lone parent | 21123 (15.9) | 10485 (49.6) | 10638 (50.4) |
| **Maternal country of birth** |  |  |  |
| UK | 115887 (87.4) | 51940 (44.8) | 63947 (55.2) |
| Outside UK | 16778 (12.7) | 8164 (48.7) | 8614 (51.3) |
| **Household tenure** |  |  |  |
| Owner occupied | 76389 (57.6) | 31657 (41.4) | 44732 (58.6) |
| Social rent | 32503 (24.5) | 18600 (57.2) | 13903 (42.8) |
| Private Rent | 23773 (17.9) | 9847 (41.4) | 13926 (58.6) |
| **Housing type** |  |  |  |
| Detached | 25378 (19.1) | 12051 (47.5) | 13327 (52.5) |
| Semi-detached | 31531 (23.8) | 11840 (37.6) | 19691 (62.5) |
| Terraced | 27108 (20.4) | 10466 (38.6) | 16642 (61.4) |
| Flat/maisonette/mobile home/temporary accommodation | 48648 (36.7) | 25747 (52.9) | 22901 (47.1) |
| **Scottish Index of Multiple Deprivation Quintile** |  |  |  |
| 1st quintile (most deprived) | 32550 (24.5) | 18737 (57.6) | 13813 (42.4) |
| 2<sup>nd</sup> | 27965 (21.1) | 12112 (43.3) | 15853 (56.7) |
| 3<sup>rd</sup> | 25890 (19.5) | 10950 (42.3) | 14940 (57.7) |
| 4<sup>th</sup> | 24531 (18.5) | 10217 (41.7) | 14314 (58.4) |
| 5th quintile (least deprived) | 21729 (16.4) | 8088 (37.2) | 13641 (62.8) |
| **Heating in home** |  |  |  |
| Gas central heating | 105058 (79.2) | 50474 (48.0) | 54584 (52.0) |
| Solid fuel heating | 1016 (0.8) | 157 (15.5) | 859 (84.6) |
| No central heating, or other | 26591 (20) | 9473 (35.6) | 17118 (64.4) |
| **Urban/rural residence** |  |  |  |
| Urban | 113004 (85.2) | 54411 (48.2) | 58593 (51.9) |
| Rural | 19661 (14.8) | 5693 (28.9) | 13698 (71.0) |

<sup>†</sup> Median postcode-level energy efficiency rating in cohort=69 (C)
<sup>a</sup>Only a small number of children had missing values. The missing category has been combined with the late term or >3,500g category respectively to avoid the risk of individual disclosure
\*There were a small number of children with a value of not applicable; their mothers may be aged less than 16 years old. The question was only asked for those aged 16 years and younger. We could not present results for this group separately due to small numbers

The differences in child, maternal, and household/neighbourhood characteristics between infants living in postcodes with below-median energy efficiency were relatively small (Table 1). The most notable difference was found according to housing tenure. The majority of infants living in socially rented housing (57.2%) lived in postcodes with high energy efficiency ratings, whereas most infants in privately rented or owner-occupied housing (58.6%) were living in postcodes with low energy efficiency ratings.

Differences in the proportion of children living in overcrowded housing were generally larger and with a reverse socio-economic pattern to energy efficiency rating. A greater proportion of pre-term infants (16.1%) and infants with born low birth weight (18.1%) lived in overcrowded households, compared to infants born at term (13.9%) or with a birthweight greater than ≥2,500g (14.9%) (Table 2). Crowding was more common among children in families with a lower socio-economic position: children born to mothers aged <20 years old were three times as likely to live in overcrowded properties compared to children born to mothers ≥30 years (30.3 cf. 9.5% of children, respectively). More than one in four children in socially rented housing lived in overcrowded households (26.3%) compared to children in privately rented (17.8%) or owner-occupied housing (7.1%).

**Table 2.**
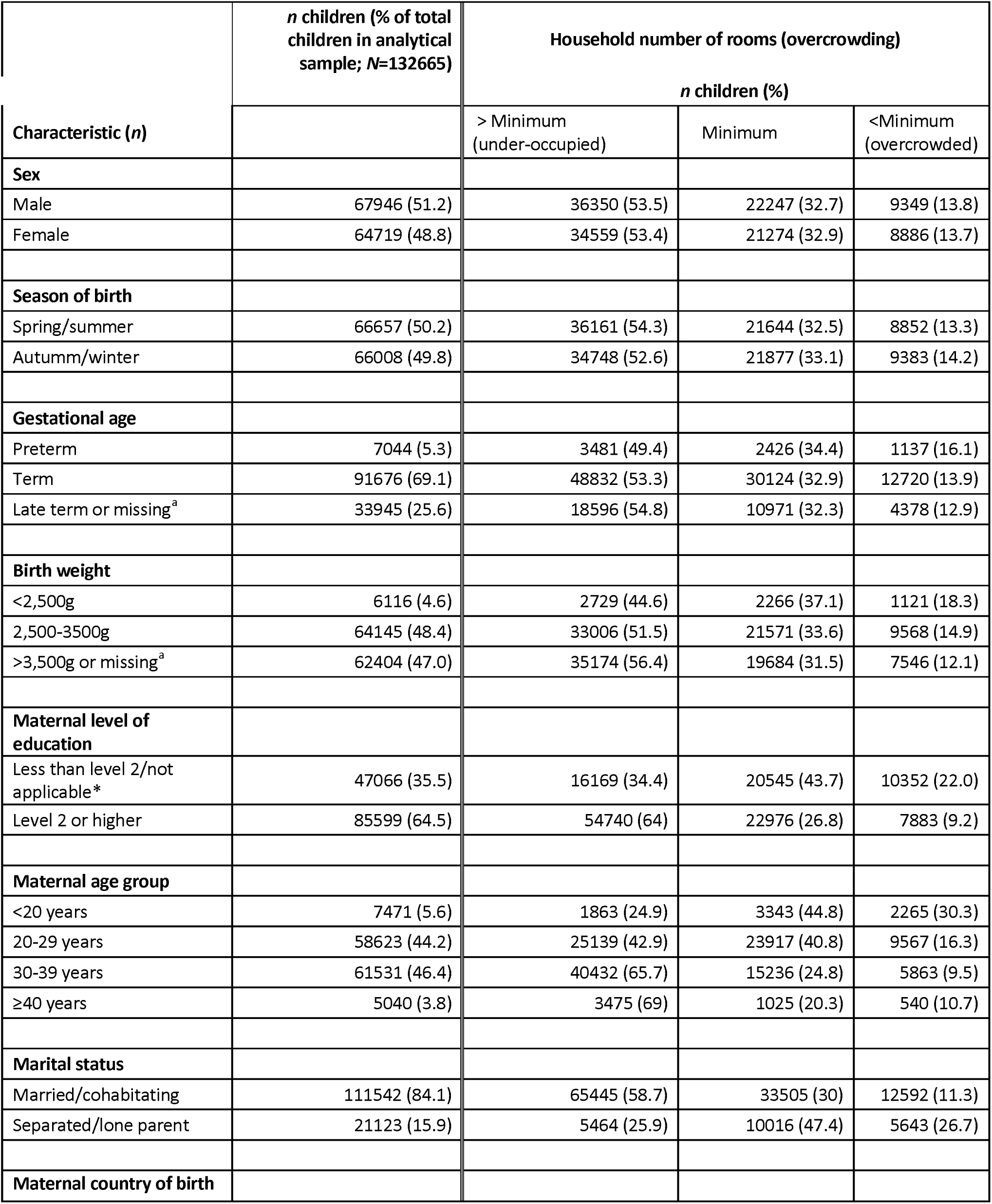
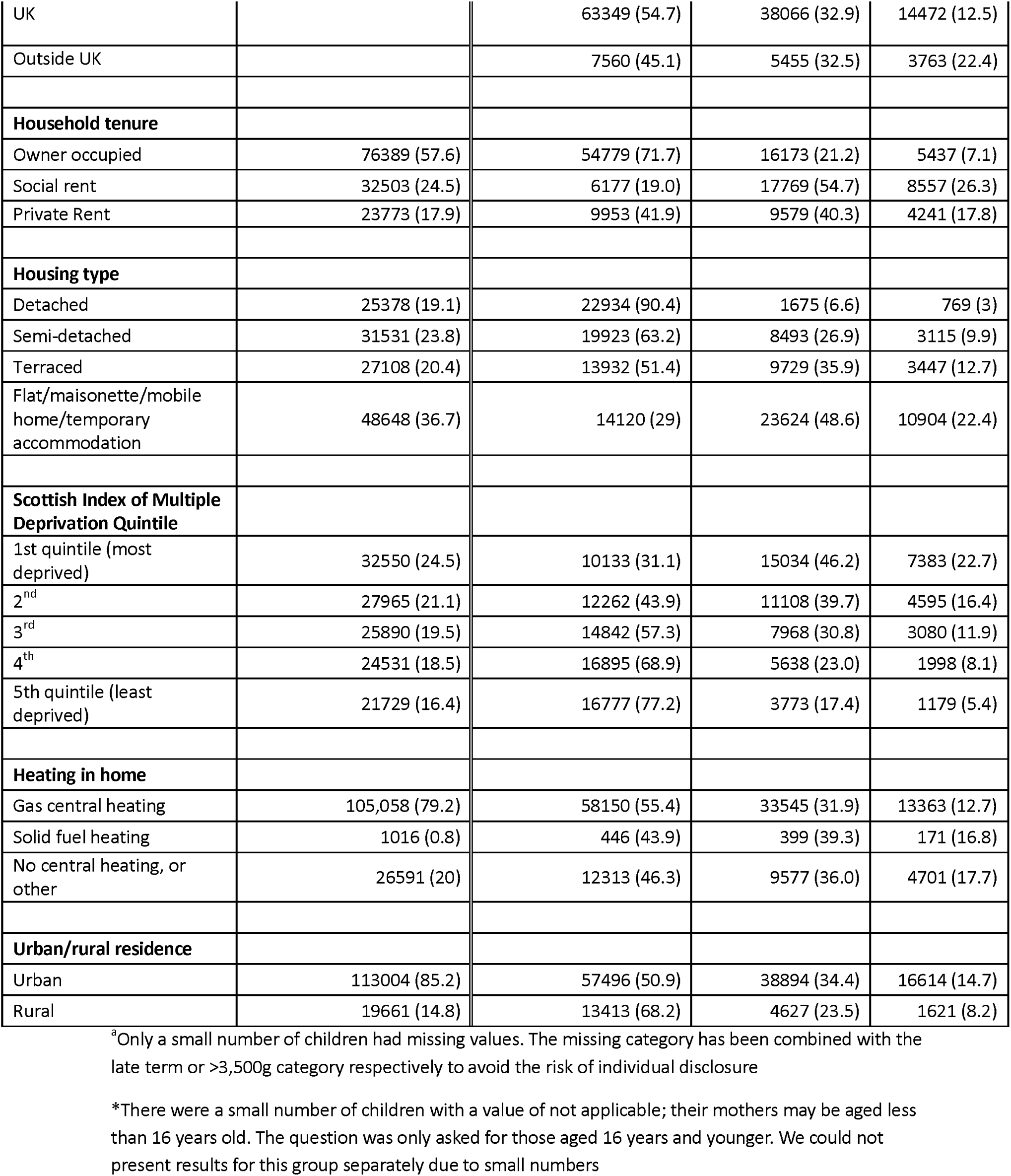
Number and proportion of infants in overcrowded households (Census 2011 room standard) according to child, household and neighbourhood characteristics, Scotland, 2010-2012. N=132,665.

Children with a birth weight <2,500g, as well as those born to mothers born outside of the UK, under 30 years old and with less than a Level 2 education, were more likely to live in postcodes with overcrowded housing and below-median energy efficiency ratings (Appendix Table A4).

In crude analyses, we found a weak association between energy efficiency ratings below the median and the odds of infant LRTI admissions (OR = 0.96, 95% CI 0.91, 1.01). We found no evidence of an association in adjusted models (Table 3).

**Table 3.** Crude and adjusted odds ratios (with 95% confidence intervals) for the association between postcode-level energy efficiency rating and household overcrowding and having one or more LRTI admissions during infancy, Scotland (2010-2012) Total children=132,665.

|  | Number of children with ≥1 LRTI admission/N (%) | Unadjusted OR (95% CI) | Adjusted OR (95% CI) |
| --- | --- | --- | --- |
| **Postcode-level energy efficiency rating\*** |  |  |  |
| High | 2480/60104 (4.1) | 1 | 1 |
| Low | 2869/72561 (4.0) | 0.96 (0.91, 1.01) | 0.99 (0.94, 1.05) |
| **Household number of rooms (overcrowding)\*\*** |  |  |  |
| > Minimum | 2538/70909 (3.6) | 1 | 1 |
| Minimum | 1991/43521 (4.6) | 1.29 (1.22, 1.37) | 1.10 (1.03, 1.17) |
| <Minimum | 820/18235 (4.5) | 1.27 (1.17, 1.37) | 1.07 (0.98, 1.17) |

†Number of observations in models =132,665 (complete case analysis)
\*adjusted for maternal age, qualifications and country of birth, household tenure, central heating type, housing type, urban/rural classification.

Compared to children living in under-occupied housing, the odds of LRTI admission were 27% higher among infants living in overcrowded housing with fewer than the minimum number of rooms (OR=1.27, 95% CI: 1.17, 1,37). The odds were also 29% higher among children living in households with the minimum number of rooms (OR=1.29, 95% CI: 1.22, 1.37). These results were substantially attenuated in models adjusted for maternal age, qualifications and country of birth, household tenure, urban/rural classification. A modest association remained for children living in overcrowded housing OR=1.07, 95% CI 0.98, 1.17), or in housing with the minimum required number of rooms (OR=1.10, 95% CI: 1.03, 1.17). There was no significant association between multiple adverse housing conditions and the odds of LRTI admission in infancy in either crude or adjusted models (Appendix Table A5).

## Discussion

Overcrowded housing was associated with an increased risk of admission to hospital with an LRTI during infancy, whereas postcode-level energy efficiency rating was not associated with the risk of infant LRTI admission. Infants in families with lower socioeconomic position, as indicated by young maternal age and living in rented, and particularly socially rented, accommodation, were more likely to live in overcrowded households. Infants born prematurely or with low birth weight were also more likely to reside in overcrowded homes, however absolute differences were small. Infants whose mothers were older, living in owner-occupied accommodation, and in less deprived areas were more likely to have lower postcode-level energy efficiency ratings.

Our use of a national linked data birth cohort allowed us to include a large proportion of children born in Scotland in the study period, minimising selection bias and loss to follow-up. Scotland’s long-standing nation-wide data linkage infrastructure allowed the integration between whole population administrative health records and environmental datasets at a fine geographical scale. Previous research in England has associated EPC variables to adult health outcomes at lower super output area (population ∼1500 individuals) level [15, 30], which are substantially larger than postcodes (population ∼40 individuals[31]). Linkage between maternal Census records and the birth cohort enabled examination of household-level overcrowding and consider several maternal and household level confounders, including multiple indicators of socio-economic position. Such detailed socio-economic variables are not available in administrative health databases.

At the time linkage was carried out for this study, 2011 Census data was the latest available for linkage to health data; future studies should update our analyses using data from the 2022 Scottish Census. Further, although linkage of EPC data at full postcode-level was a major advancement compared to previous work, the ideal would have been linkage at individual address level. This was not possible at the time in Scotland, however unique property reference numbers (UPRNs; individual identifiers of each addressable location in the UK), have since been linked to the Scottish CHI register, including historic addresses, making it possible to link EPC and other property or environmental data to health data over time. This linkage methodology will be implemented Scotland-wide for the Homes, Heat and Healthy Kids study[32].

EPCs were not available for all properties in a postcode. The average ratio of EPCs per 2011 property count per postcode was 0.55, but the range was 0.02 and 1.3 (the ratio is>1 for postcodes where new properties had been built since the 2011 Census, and the new properties had at least one EPC). Nevertheless, we considered postcode-level energy efficiency to be a reasonable indicator of exposure to energy efficient homes across a population. Further, we linked to the most recent EPC available within a postcode using the most recent version of the EPC register at time of linkage-2021. This was to ensure as many EPCs as possible for each postcode; and we considered some misclassification would still occur since updates of the EPC register are not mandatory unless a property is bought or sold. We therefore assumed that the latest energy efficiency ratings on the 2021 register reflected energy efficiency ratings during 2010-2012. Median energy efficiency in Scotland increased from a median rating of 62 to 69 between 2010 and 2019 across all households, with retrofitting more common in less energy efficient dwellings [33]. This may have resulted in an underestimation of the OR between energy efficiency and LRTI admissions.

We did not allow for residential mobility and therefore assumed that exposure to overcrowding did not change throughout infancy. Although residential mobility rates are not published for infants separately, 16% of children aged 1-2 years old taking part in the Growing Up In Scotland cohort study moved address at least once, and moving was more common for younger mothers and children living in rented accommodation[34]. We reduced exposure misclassification bias arising from residential mobility by limiting follow-up to the end of infancy and only linking to maternal Census records for children who were born a year before or after the Census year.

Previous studies [10, 35] have identified household overcrowding as a risk factor for respiratory infections in children, although many different definitions of overcrowding have been used. Overcrowding facilitates the transmission of respiratory infections[36], and is associated with a higher probability of damp and mould, themselves risk factors for more severe respiratory symptoms[37]. We found that infants living in crowded households, as well as those living in households where the number of rooms was equal to the minimum required, had increased odds of LRTI admissions in infancy compared to infants living in under-occupied households. The elevated odds among children in households where the number of rooms was equal to the standard could be due to unadjusted confounding. Alternatively, meeting the Scottish Census minimum rooms standard may not preclude levels of crowding associated with an increase in risk of severe LRTIa in infants. The poor suitability of some overcrowding indicators for children’s health outcomes has also been raised by others[38]. The 2022 Census uses the number of bedrooms, rather than the number of rooms to calculate over or under-occupancy of a property[39].

We found that the proportion of infants living in overcrowded housing (13.7%) was higher than for the population of Scotland overall (9% in 2011)[40], mirroring data from England and Wales that families with dependent children are more likely to live in overcrowded housing than older people[41]. The Scottish Housing Conditions Survey, which like the 2022 Census measures overcrowding in relation to the number of bedrooms, shows that overcrowding has remained relatively constant since 2012[25]. Our findings suggest that overcrowding, which remains a persistent housing issue for families with young children, may be an important determinant of infant respiratory health and contribute to inequalities in LRTI admissions.

Lower energy efficiency, unlike overcrowding, was correlated with higher socio-economic position, and not associated with LRTI risk in children. Energy efficiency of dwellings is related to the heating system, building fabric and retrofit history. Houses and older homes are therefore generally less energy efficient compared to flats/apartments and new- builds, yet are housing families with higher socio-economic position. Although we adjusted for various measures of socio-economic position, our results for energy efficiency and its link to LRTI admission odds may be due to unadjusted confounding. In the UK, including Scotland, retrofitting rates of homes to improve energy efficiency has been consistently higher among socially rented, rather than privately rented or owner-occupied owned homes[42], reflecting sustained investment in improving social housing[43, 44]. However, we previously showed, using the same cohort[22], no difference in LRTI admission rates in children<2 years old between children living in private vs social rented homes.

Sharpe et al.[15] found a positive relationship between higher rates of energy efficiency improvements in homes and respiratory hospital admissions in adults in England at small-area level. They hypothesised that more energy efficient buildings may have worse ventilation thereby trapping indoor air pollution and enhancing transmission of respiratory pathogens. We could not replicate these findings among infants in Scotland. However, our EPC data was at postcode-level; future studies with address-level data on EPC would be able to test the association between energy efficient homes and respiratory admissions more robustly.

Our housing measures captured different dimensions of housing quality. While overcrowding was more common among children living in families with a lower socio- economic position and was associated with an increased risk of LRTI admission during infancy, postcode-level energy efficiency showed a different socioeconomic pattern and was not associated with LRTI admissions. Ensuring that families with young children have access to housing with living space proportionate to family size may help reduce the burden of LRTIs in infancy.

## Ethical considerations

This study was approved by the NHS Scotland Public Benefit and Privacy Panel for Health and Social Care (HSC-PBPP) and the Statistics Public Benefit and Privacy Panel, reference 1819-0049, and the University of Edinburgh School of Geosciences Ethics Committee, reference 2020-401.

## Data Availability

The data used in this paper are not openly available and cannot be reshared by the authors. Access can be requested via the Electronic Data Research and Innovation Service (eDRIS) Scotland: https://publichealthscotland.scot/resources-and-tools/health-intelligence-and-data-management/electronic-data-research-and-innovation-service-edris/overview/what-is-edris/

## Acknowledgements

We acknowledge the support of the Electronic Data Research and Innovation Service (eDRIS) team at Public Health Scotland and the Data Access Team at National Records for Scotland without whom this study would not have been possible. We are particularly grateful to Hannah Mackenzie, Diane Rennie and Liam Cavin. This work uses data provided by patients and collected by the NHS as part of their care and support.

## Author contributions

Conceptualization: CH, PH, BDS; Data curation and formal analysis CH, JT, PH, BDS; Funding acquisition: PH, BDS, SC, CD, JT; Methodology: CH, PH, BDS, JT; Supervision: PH, SH and BDS; Writing-original draft: PH and CH; Writing-review and editing: PH, AR, CH, MR, BDS, SH, SC, PS, CD, OS, JT

## residential crowding and lower respiratory tract infection admissions in infants: national birth cohort study in Scotland – Appendix A

**Table A1.** Data sources used to develop cohort, with variables obtained from each dataset, Scotland, 2010-2012.

| Type | Source | Source Description | Variables of interest |
| --- | --- | --- | --- |
| **Birth Records** | National Records of Scotland (NRS) vital statistics records. | Registrations of births, and deaths occurring in the first year of life. | Week of birth<br>Year of birth<br>Baby sex<br>Area of residence (Urban/rural)*<br>Date of death |
|  | Scottish Birth Record | Patient level record of neonatal care, from 2002 onwards. | Week of birth |
| **Postcode Records** | Community Health Index Database | Longitudinal postcode histories | Postcode (de-identified)<br>Date arriving and leaving postcode<br>Date leaving Scotland |
| **Maternity/delivery Records** | Scottish Morbidity Record 02 (SMR-02) | Patient level longitudinal hospital obstetric records for the mother | Birth weight<br>Estimated gestational age:<br>Maternal age |
| **Hospital Admission Records** | Scottish Morbidity Record 01 (SMR-01) | Patient level hospital Longitudinal hospital admission records for the child. | Admission date<br>Discharge date<br>Admission Type<br>Main and additional diagnoses (ICD-10) |
| **Maternal Census records (2011)** | Decennial Census records (mother) | Resident and household social and demographic information. | Maternal highest qualification level<br>Housing tenure<br>Accommodation type<br>Occupancy rating<br>Heating Type |
| Energy Performance Certificates (EPCs) | Scottish Government | Dwelling-level information on housing characteristics<br>.<br>Aggregated to postcode-level for linkage in this study | Median Energy Efficiency rating |

\*Area of residence was derived from the *UR8* variable. We assigned the classification closest to the child's birth year dichotomised it into urban/rural.

**Table A2.**
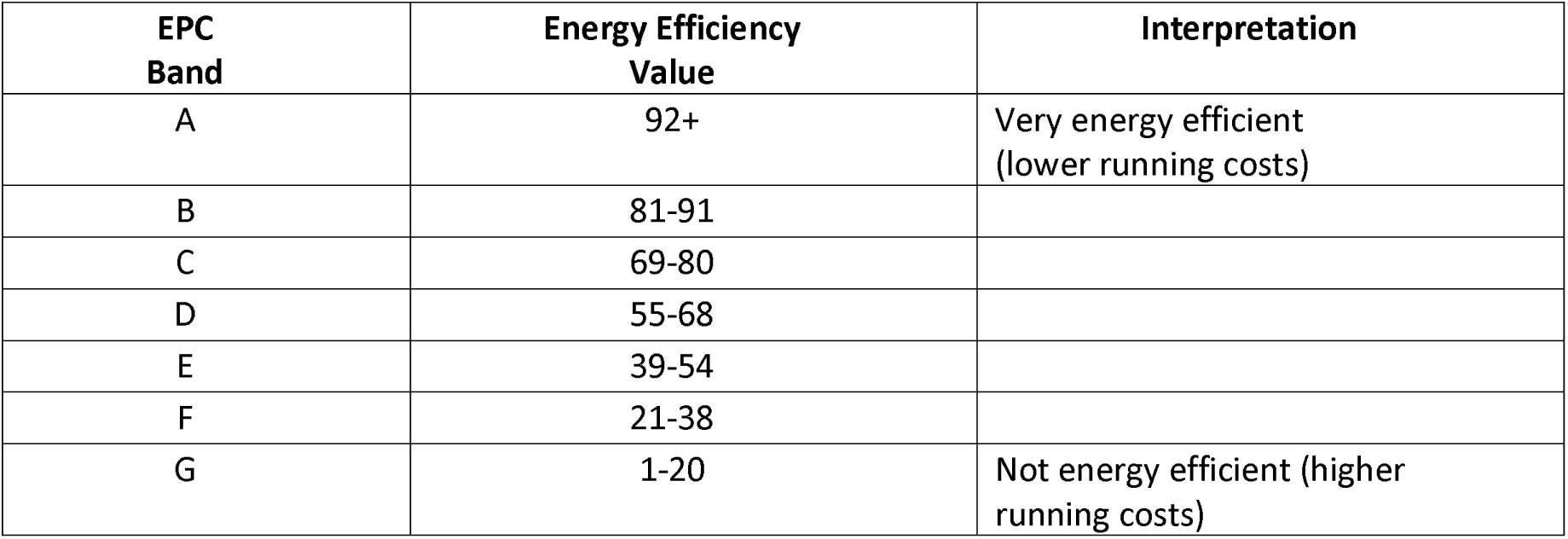
Energy Performance Certificate ratings, energy efficiency values, and interpretations.

**Table A3.**
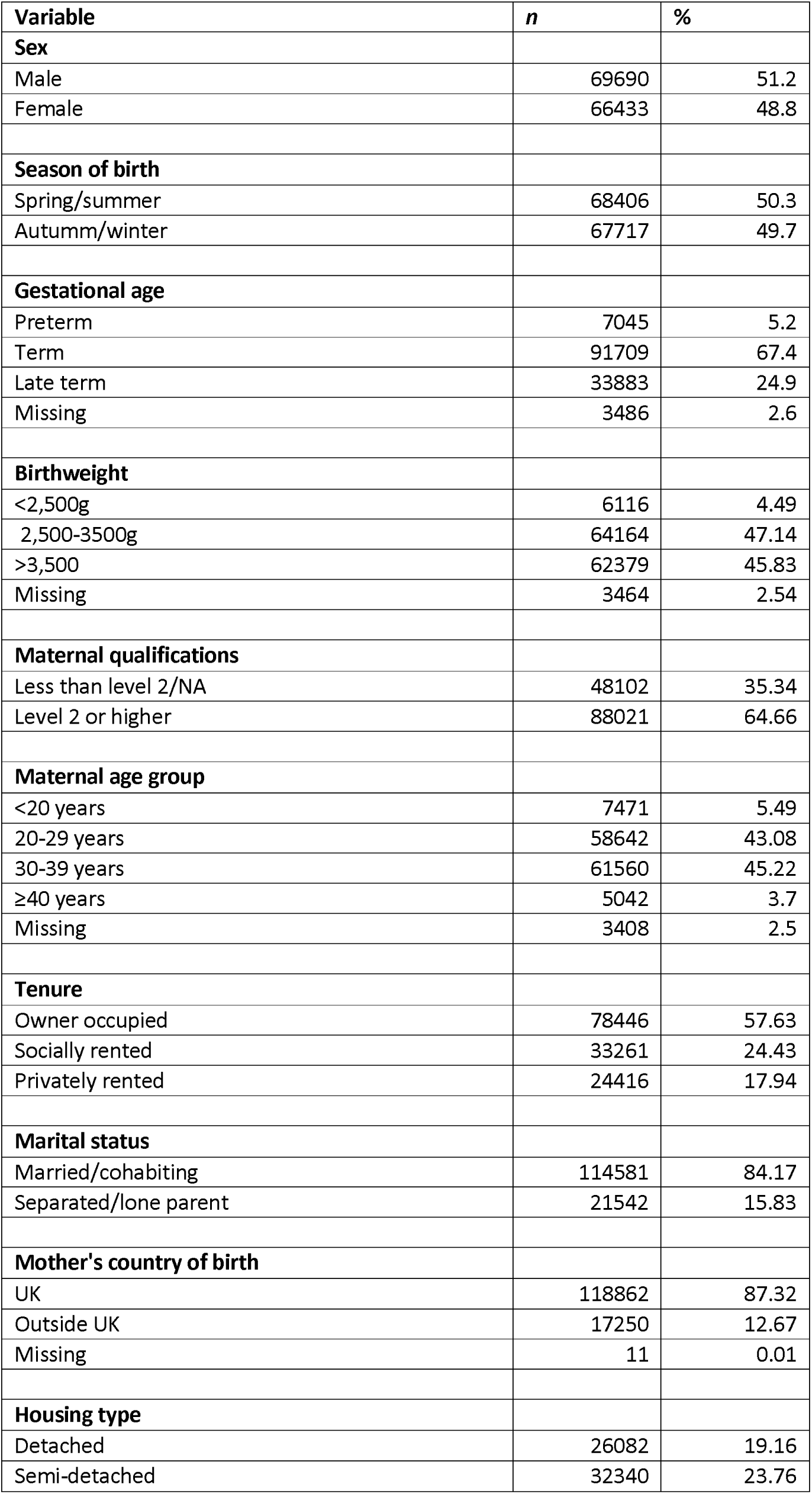
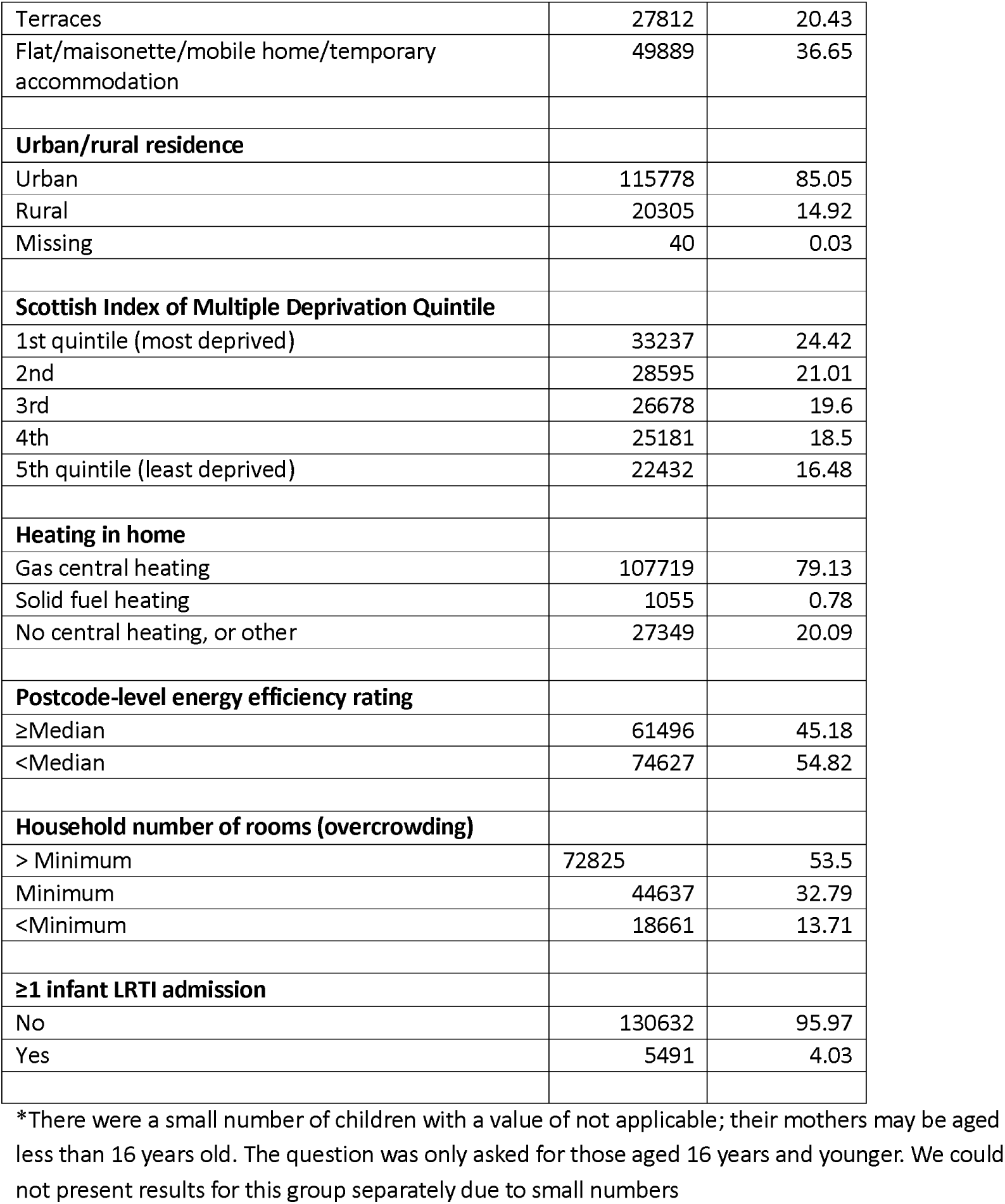
Distribution of variables of interest among infants in the full cohort (including children with missing data on variables used for statistical modelling), Scotland, 2010-2012. Total N=136,123.

**Table A4.**
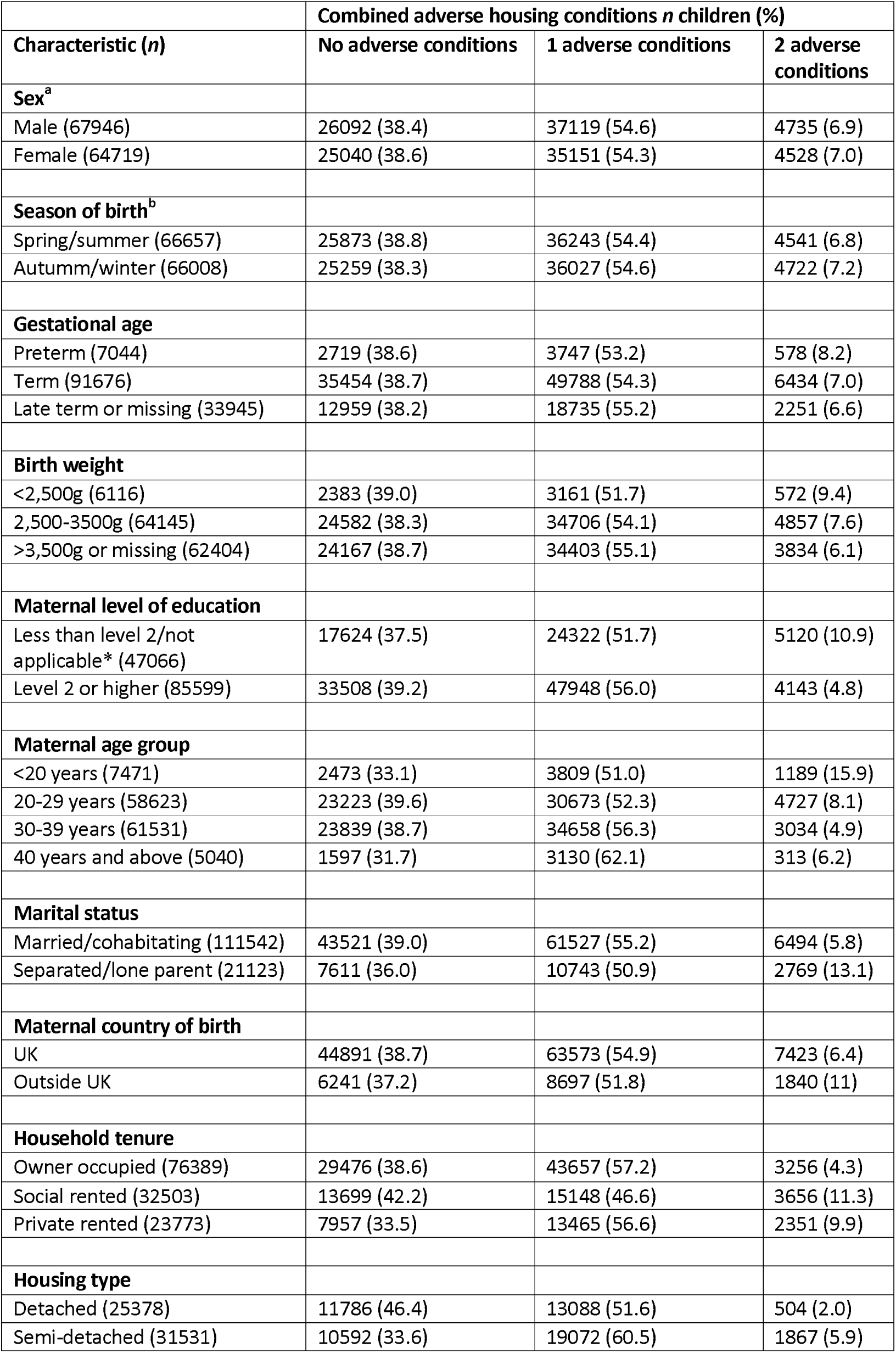
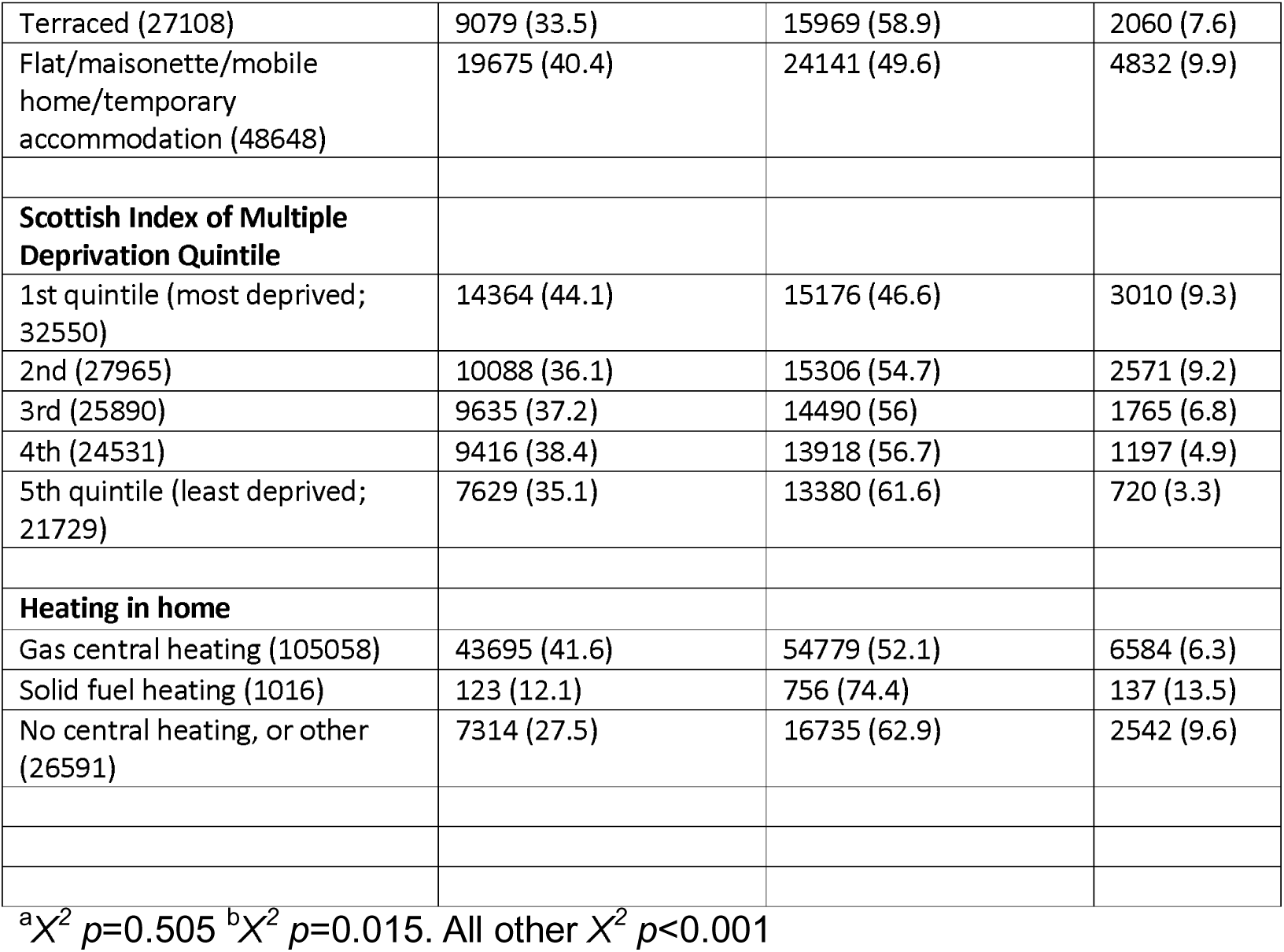
Distribution of combined adverse housing condition variable according to child, family, household and neighbourhood characteristics, Scotland, 2010-2012 (*N*=132,665).

**Table A5.** Number and % of children with one or more LRTI admissions during infancy, with adjusted and unadjusted odds ratios (ORs), with 95% confidence intervals (95% CIs), according to the number of adverse housing conditions^†^.

| **Number of adverse housing conditions\*** | **number of children/N (%)** | **unadjusted OR (95% CI)** | **adjusted OR (95% CI)** |
| --- | --- | --- | --- |
| No adverse conditions | 2063/51132 (4) | 1 | 1 |
| 1 adverse conditions | 2883/72270 (4) | 0.99 (0.93 ,1.05) | 1.01 (0.95 ,1.07) |
| 2 adverse conditions | 403/9263 (4.4) | 1.08 (0.97 ,1.21) | 1.01 (0.91 ,1.13) |

<sup>†</sup>Number of observations in models =132,665 (complete case analysis)
\*adjusted for maternal age, qualifications and country of birth, household tenure, central heating type, housing type, urban/rural classification.

**Figure A1.**
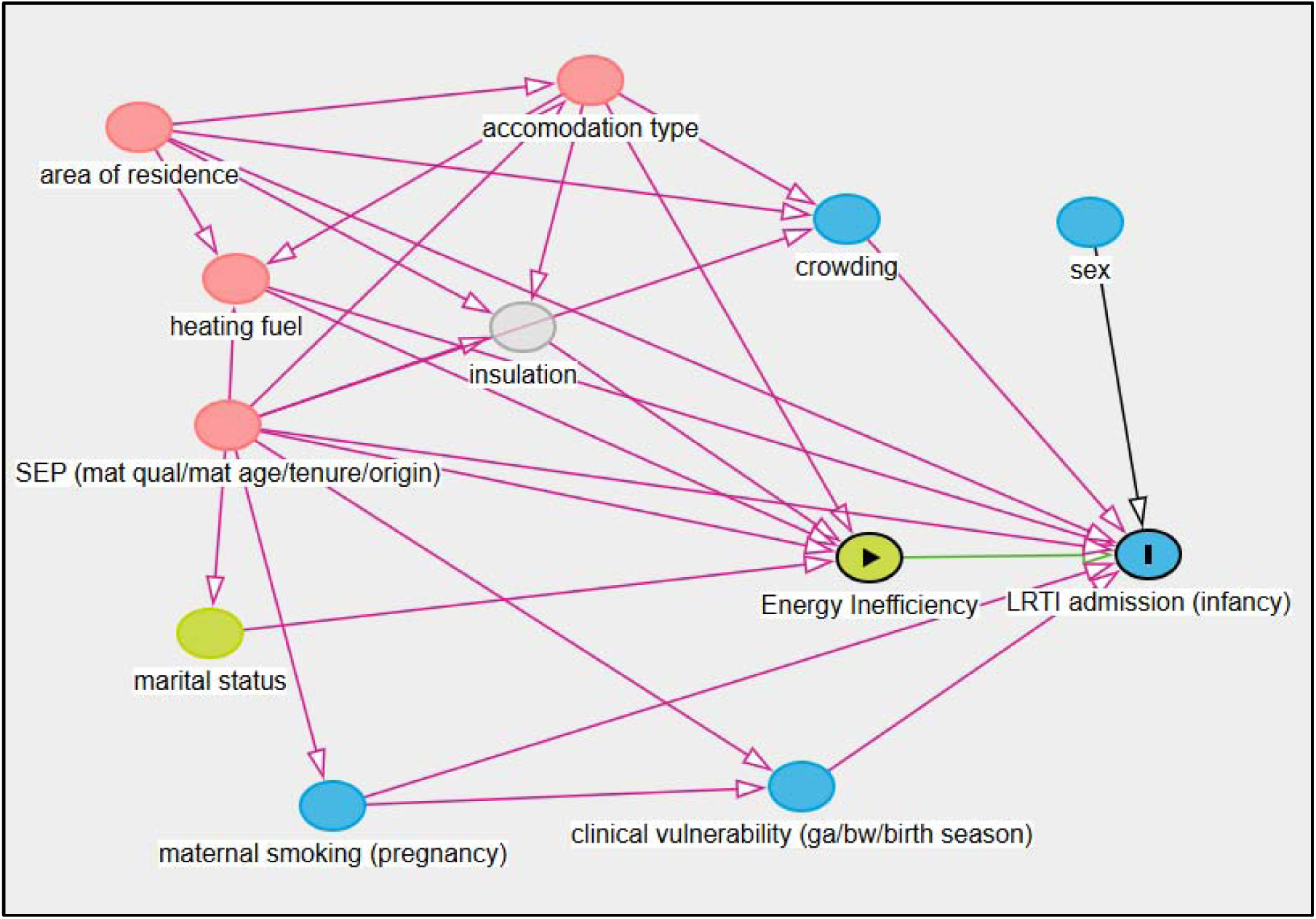
Directed acyclic graph illustrating the assumed relationships between energy efficiency, infant LRTI admission, and potential confounders.

Pink nodes indicate variables that are ancestors of both the exposure and the outcome and were identified as the minimal sufficient adjustment set for estimating the total effect. These included accommodation type, heating/fuel type, socio-economic position (maternal qualifications, tenure, maternal age, and country of birth/origin), and area of residence. Red paths indicate open backdoor paths, representing potential biasing pathways that should be blocked through adjustment

The minimal sufficient adjustment sets, identified for estimating the total effect of energy efficiency on infant LRTI admission included accommodation type, heating/fuel type, socioeconomic position (maternal qualifications/tenure/maternal age/country of birth [origin]), and urban/rural classification (area of residence)

**Figure A2.**
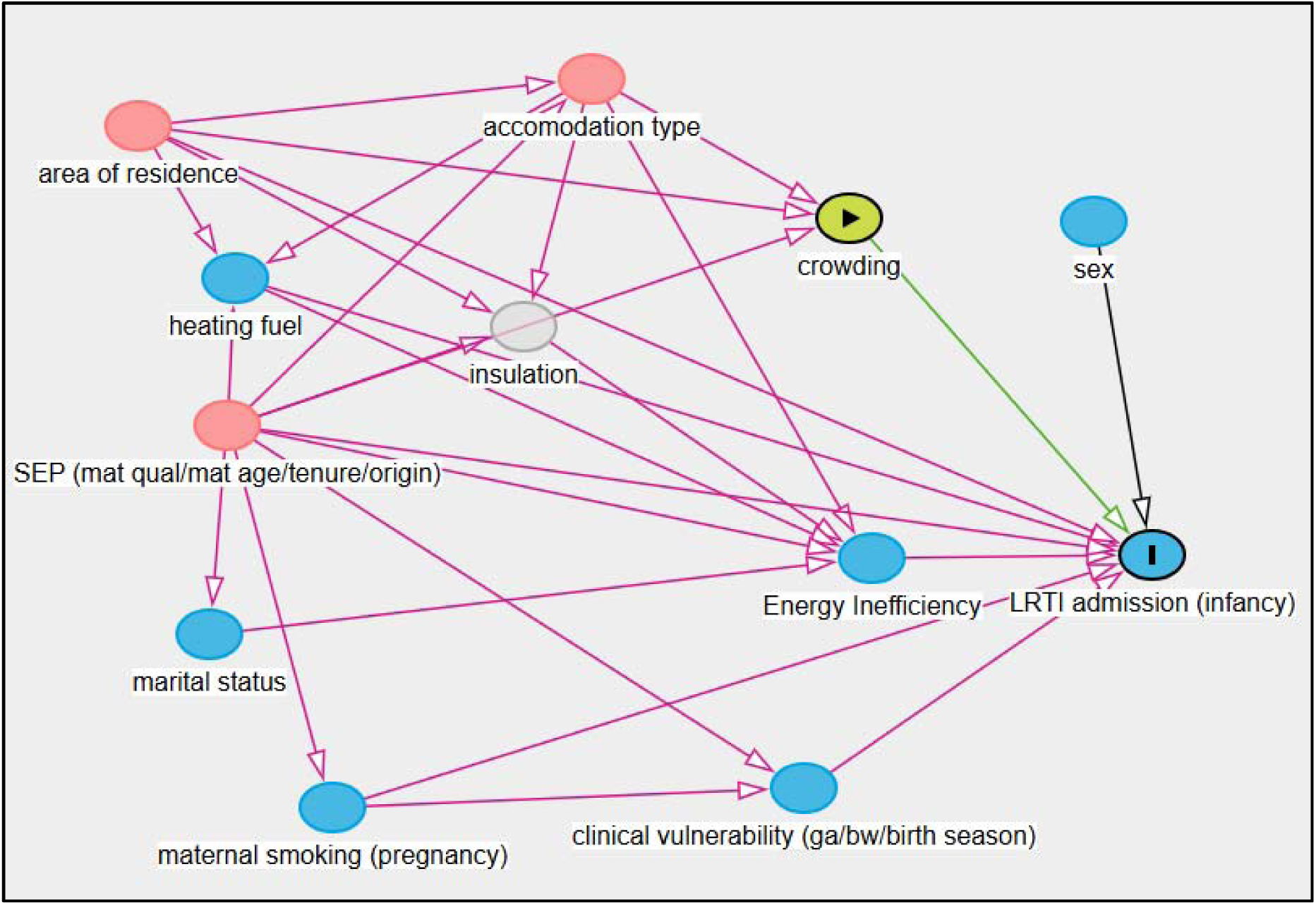
Directed acyclic graph of the presumed relationships between residential overcrowding and infant LRTI admission.

Pink nodes indicate variables that are ancestors of both the exposure and the outcome and were identified as the minimal sufficient adjustment set for estimating the total effect. These included accommodation type, heating/fuel type, socio-economic position (maternal qualifications, tenure, maternal age, and country of birth/origin), and area of residence. Red paths indicate open backdoor paths, representing potential biasing pathways that should be blocked through adjustment

The minimal sufficient adjustment sets, identified for estimating the total effect of overcrowding on infant LRTI admission included socioeconomic position (maternal qualifications/tenure/maternal age/country of birth [origin]), and urban/rural classification (area of residence)

